# Patterns of modifiable lifestyle, behavioural and health-related risk factors and their associations with incident dementia in UK Biobank

**DOI:** 10.64898/2026.09.24.26363838

**Authors:** Yamato Uejima, Catherine M Calvin, Elżbieta Kuźma, Thomas J. Littlejohns

## Abstract

**INTRODUCTION:** Individual lifestyle, behavioural and health-related factors are associated with the risk of developing dementia. However, the co-occurrence of these factors, which are likely to form distinct patterns, in relation to dementia risk has been understudied.

**METHODS:** The sample included 151,832 UK Biobank participants aged 60-69 years. Latent class analysis was used to identify patterns among participants with two or more of the following risk factors: high alcohol consumption, high body weight, depression, diabetes, poor diet, hypertension, physical inactivity, visual impairment, hearing impairment, sleep disturbance, smoking, social isolation, traumatic brain injury, and vitamin D deficiency. Multivariate Cox proportional hazards models were used to determine the associations between identified risk factor patterns and incident dementia ascertained through hospital and death records.

**RESULTS:** Over a median follow-up of 13 years, 5,304 participants developed dementia. Six risk factor classes were identified. Compared to participants with zero or one risk factor, all classes were associated with dementia risk: ‘multiple risk factors’ (hazard ratio [HR] 2.38, 95% confidence interval [CI] 2.11-2.68), ‘hearing loss and unhealthy lifestyle’ (HR=1.37, 95% CI 1.22-1.55), ‘sleep and psychosocial factors’ (HR=1.34, 95% CI 1.20-1.49), ‘poor cardiometabolic health’ (HR=1.32, 95% CI 1.19-1.46), ‘heavy drinking and unhealthy lifestyle’ (HR=1.20, 95% CI 1.07-1.35), and ‘vitamin D deficiency, sedentary and psychosocial factors’ (HR=1.18, 95% CI 1.03-1.35).

**DISCUSSION:** We identified risk factor patterns that were differentially associated with dementia risk. These findings suggest that accounting for the co-occurrence of risk factors might be an effective approach for risk stratification and developing effective dementia prevention strategies.

**Highlights:**

- There is limited research on identifying clusters of lifestyle, behavioural, and health-related factors linked to dementia risk.
- Six distinct classes of modifiable dementia risk factor patterns were identified in UK Biobank participants.
- All classes were associated with higher dementia risk, strongest for those with multiple coexisting risk factors.
- The findings support the importance of multi-domain approaches to dementia prevention.

## Introduction

Over 55 million people worldwide live with dementia, a number projected to increase to 150 million by 2050 due to population aging. This is cause for concern, as dementia is currently the seventh leading cause of death globally and a major source of disability among older adults.^1^ Promisingly, the 2024 Lancet Commission estimated that the onset of 45% of all dementia cases could be delayed or prevented by addressing a range of lifestyle and health-related modifiable risk factors.^2^ Dementia prevention typically focuses on the role of individual risk factors; however, unhealthy lifestyle behaviours (i.e., smoking and high alcohol consumption) and health conditions (i.e., diabetes and hypertension) rarely act in isolation and commonly cooccur in the same individual.

Several studies have shown that a cumulative increase in the number of risk factors is associated with dementia risk. ^3^ However, this approach assumes that each factor contributes equally to dementia risk and does not account for potentially distinct patterns of cooccurring factors, which might have even stronger associations with dementia risk. Studies that have identified clusters in relation to dementia risk have derived clusters for a small selection of either lifestyle behaviours^4–6^ or cardiometabolic factors.^7^ For example, Dingle and colleagues reported that physically inactive individuals with low social engagement had a greater risk of dementia than smokers and alcohol drinkers with high social engagement.^4^ These findings suggest that sociability could moderate the associations between other factors and dementia risk. A more recent study extended this approach by examining a broader set of modifiable risk factors; Xiong and colleagues identified three sex-specific profiles (cardiometabolic, substance use-related, and low risk) and reported that these profiles were differentially associated with dementia incidence.^8^ Identifying patterns across the full range of lifestyle, behavioural, and health-related factors (including vascular, metabolic, mental health and sensory systems) linked to dementia risk could provide novel insights to inform dementia prevention strategies, particularly as almost half of dementia prevention trials are conducting multidomain interventions, with an increasing number of trials applying personalised, targeted approaches on the basis of risk factor profiles.^9, 10^

To achieve this goal, it is necessary to utilise a dataset with a large sample size, detailed data collection and sufficiently long follow-up to provide confidence that preclinical dementia pathology does not influence risk factor measures. In the present study, we identified clusters of diverse risk factors, selected a priori from a comprehensive umbrella review, in a population-based cohort of 150,000 participants and examined their associations with dementia risk over a 15-year follow-up period.

## Methods

### UK Biobank population

The UK Biobank is a population-based prospective cohort study of 502,416 women and men aged 40--69 years who were recruited between 2006 and 2010 across England, Scotland and Wales.^11^ All participants attended a baseline assessment centre where they completed touchscreen questionnaires and a verbal interview, underwent physical examinations and provided biological measures. All participants provided electronically signed informed consent. The UK Biobank received ethical approval from the National Health Service North West Centre for Research Ethics Committee (Ref: 11/NW/0382).

To ensure that the study population was restricted to individuals at risk of developing dementia over the follow-up period, 284,907 participants aged less than 60 years at baseline were excluded. Of the remaining 217,509 participants, 167 with prevalent self-reported or hospital-diagnosed dementia were excluded, yielding 217,342 eligible participants.

### Risk factor exposures

Exposures were selected from a 2024 umbrella review published by Jones and colleagues^12^, which synthesised 45 systematic reviews with 212 meta-analyses on modifiable risk factors for dementia. The review identified 14 factors significantly associated with dementia risk: high alcohol consumption, high body weight, depression, diabetes mellitus, poor diet, hypertension, low education, physical inactivity, sensory loss, sleep disturbance, smoking, social isolation, traumatic brain injury, and vitamin D deficiency. All risk factors were measured in the UK Biobank, and a detailed description of how each was defined in the current study is provided in **Table S1**. Education was excluded from the risk factors because it is an early-life factor that, unlike the other 14 risk factors, does not represent a contemporaneous modifiable exposure. Including education alongside current lifestyle and health behaviours could conflate different temporal windows of exposure and complicate interpretation. Education was instead adjusted for as a covariate in all regression analyses. Furthermore, we separated ‘sensory loss’ into hearing impairment and visual impairment. The population was restricted to participants with complete data for all 14 risk factors to ensure that missing data did not bias the identification of different clusters.

### Incident all-cause dementia

All-cause dementia was determined via hospital inpatient and death registry records. Hospital inpatient records were obtained from the Hospital Episode Statistics (HES) for England, the Scottish Morbidity Records (SMRs) for Scotland, and the Patient Episode Database (PEDW) for Wales. Death registry records were obtained from NHS England for England and Wales and the NHS Central Register, National Records of Scotland for Scotland. Primary and secondary hospital diagnoses and underlying and contributory causes of death were recorded via the International Classification of Diseases (ICD) coding system. The ICD codes used to ascertain dementia cases in the current study were previously selected and validated by the UK Biobank outcome adjudication group (**Table S2**).^13^

### Covariates

Covariates were selected on the basis of a previous study^14^ using the UK Biobank and have been identified as important confounders owing to the associations between various risk factors and dementia. The covariates included age in years at baseline and sex (female, male). Ethnic background (white, nonwhite) and highest educational attainment (primary, secondary, postsecondary nontertiary, tertiary) were self-reported during the touchscreen questionnaire. Socioeconomic status (in quintiles) was based on the Townsend deprivation score^15^, which is a proxy for material socioeconomic deprivation and assigned to each study participant on the basis of their residential postal code at baseline. The apolipoprotein (APOE)-ε4 carrier status (noncarrier or carrier) was determined via the rs429358 and rs7412 single nucleotide polymorphisms, which were directly genotyped on the UK Biobank arrays from blood samples.^16^ Given the minimal amount of missing data (less than four percent for any variable) and to preserve statistical power, participants with any missing covariate data (ethnicity, education, socioeconomic status, and APOE-ε4 carrier status) were assigned to a separate category for each categorical variable.^17^

### Statistical analysis

Latent class analysis (LCA) was employed to derive classes of selected risk factor patterns among participants with two or more risk factors. Consistent with the prior literature,^14^ a random sample of 80% of participants with two or more risk factors (training sample) was used to determine the optimal number of classes and subsequently estimate the association of risk factor classes with dementia risk compared with a reference category of zero or one risk factor. Output statistics were generated for multiple LCA models, ranging from one to ten class solutions. A six-class model was selected on the basis of model fit statistics, which included a relatively low Bayesian information criterion (BIC), sample size-adjusted BIC, and likelihood ratio statistics (**Figure S1**). The selection of six classes was further supported by higher class separation (entropy), with little gains in entropy and loss of clinical interpretability when additional classes were added. The participants were assigned to classes with the highest estimated posterior probability, indicating their most likely latent class. The expected prevalence of each risk factor among all the participants with two or more risk factors was computed to estimate the observed and expected prevalence ratios (O/E).

Each class was characterised by risk factor patterns with higher observed prevalence, excluding those with an O/E of less than one (i.e., the observed prevalence was less than the expected prevalence). When characterising each class, in addition to the higher level of observed prevalence, the O/E level was considered to account for varying expected prevalence levels of each risk factor. To validate the class solutions, the remaining 20% of participants with two or more risk factors (test sample) were subjected to LCA, with the number of classes fixed to match the optimal number determined from the training set.

Cox proportional hazards models adjusted for age, sex, ethnicity, education, socioeconomic status, and APOE-ε4 carrier status were used to estimate the associations between 1) the number of risk factors and 2) risk classes with incident all-cause dementia. For all analyses, participants with 0–1 risk factors were used as the reference group. To ensure comparability across training and test datasets, the same 80/20 split was also applied to the zero or one risk factor group. This yielded a training dataset of 121,468 participants (98,506 with ≥2 risk factors, 22,962 with 0–1 risk factors) and a test dataset of 30,364 participants (24,624 with ≥2 risk factors, 5,740 with 0–1 risk factors). While training dataset was used for the main analysis, test dataset was used to validate the latent class and the associations between these classes and dementia risk in both samples were compared. Person-years were calculated from the date of baseline assessment until the date of first dementia diagnosis, death, loss to follow-up, or the end of follow-up, whichever occurred first. The end of follow-up was based on the availability of electronic health record data, which were censored on October 31, 2022, for England; August 31, 2022, for Scotland; and May 31, 2022, for Wales. All models were assessed for the proportional hazards assumption via statistical tests and visual inspection of Schoenfeld residuals; no significant violations were observed. While conventional 95% CIs are reported in the text, floating absolute risks were used in the forest plots to estimate group-specific 95% CIs for all categories, including the reference group. ^18^ This approach avoids the arbitrary assumption that the reference group has no uncertainty and facilitates graphical comparisons between any two groups, rather than only comparisons with the reference group.

To investigate the potential role of reverse causation whereby prodromal dementia affects risk factors in the short term, a sensitivity analysis was conducted by stratifying the follow-up time into two periods (<10 years and ≥10 years). Additionally, effect modification was investigated by incorporating interaction terms between each risk factor class and 1) APOE-ε4 carrier status and 2) sex, and subgroup effects were estimated.

Several sensitivity analyses were conducted to assess the robustness of the main findings. First, attained age was used as the time scale. Second, participants with prevalent cardiovascular disease (CVD) at baseline — defined as self-reported history of coronary heart disease, heart failure, cerebrovascular disease, or aortic and peripheral arterial disease — were excluded, and Cox models were re-estimated. Third, the reference group was restricted to participants with zero risk factors only, with participants with one risk factor retained as a separate comparison category.

P values were 2-sided, and the type I error rate for statistical significance was set at 0.05. All analyses were conducted in R (version 4.2.2); the poLCA package^19^ was used for LCA.

## Results

A total of 151,832 participants were included in the final sample (**Figure S2**). Participants with two or more risk factors were more likely to be male and have lower educational qualifications and lower socioeconomic status than those with zero or one risk factor (**Table 1**). The prevalence of each of the 14 risk factors among the total participants ranged from 0.4% for traumatic brain injury to 49% for low vitamin D.

**Table 1.** Descriptive statistics by number of risk factors (total sample)

| Characteristics | Total<br>N=151,832 | 0-1 risk factor<br>N=28,702 | ≥2 risk factors<br>N=123,130 | P-value |
| --- | --- | --- | --- | --- |
| Age in years, mean (SD) | 64.1 (2.8) | 63.9 (2.8) | 64.1 (2.9) | <0.001 |
| Sex |  |  |  | <0.001 |
| Female | 74,757 (49.2%) | 17,704 (61.7%) | 57,053 (46.3%) |  |
| Male | 77,075 (50.8%) | 10,998 (38.3%) | 66,077 (53.7%) |  |
| Ethnicity |  |  |  | <0.001 |
| Non-White | 3,585 (2.4%) | 350 (1.2%) | 3,235 (2.6%) |  |
| White | 147,807 (97.3%) | 28,292 (98.6%) | 119,515 (97.1%) |  |
| Missing | 440 (0.3%) | 60 (0.2%) | 380 (0.3%) |  |
| Education |  |  |  | <0.001 |
| Primary | 35,742 (23.5%) | 5,673 (19.8%) | 30,069 (24.4%) |  |
| Secondary | 49,554 (32.6%) | 10,103 (35.2%) | 39,451 (32.0%) |  |
| Post-secondary non-<br>tertiary | 22,006 (14.5%) | 3,949 (13.8%) | 18,057 (14.7%) |  |
| Tertiary | 43,296 (28.5%) | 8,739 (30.4%) | 34,557 (28.1%) |  |
| Missing | 1,234 (0.8%) | 238 (0.8%) | 996 (0.8%) |  |
| Socioeconomic Status,<br>quintile |  |  |  | <0.001 |
| Least deprived | 31,686 (20.9%) | 7,165 (25.0%) | 24,521 (19.9%) |  |
| Second least deprived | 31,299 (20.6%) | 6,589 (23.0%) | 24,710 (20.1%) |  |
| Middle | 30,694 (20.2%) | 6,157 (21.5%) | 24,537 (19.9%) |  |
| Second most deprived | 30,145 (19.9%) | 5,235 (18.2%) | 24,910 (20.2%) |  |
| Most deprived | 27,882 (18.4%) | 3,537 (12.3%) | 24,345 (19.8%) |  |
| Missing | 126 (0.1%) | 19 (0.1%) | 107 (0.1%) |  |
| APOE-ε4 carrier status |  |  |  | 0.071 |
| Non-carrier | 108,646 (71.6%) | 20,426 (71.2%) | 88,220 (71.6%) |  |
| Carrier | 37,768 (24.9%) | 7,194 (25.1%) | 30,574 (24.8%) |  |
| Missing | 5,418 (3.6%) | 1,082 (3.8%) | 4,336 (3.5%) |  |
| Alcohol drink |  |  |  | <0.001 |
| 1 to 20 units per week | 106,285 (70.0%) | 25,178 (87.7%) | 81,107 (65.9%) |  |
| Non-drinker | 12,157 (8.0%) | 1,994 (6.9%) | 10,163 (8.3%) |  |
| 21 units and more per week | 33,390 (22.0%) | 1,530 (5.3%) | 31,860 (25.9%) |  |
| High body mass index, $\geq 30$ kg/m<sup>2</sup>. | 36,235 (23.9%) | 783 (2.7%) | 35,452 (28.8%) | <0.001 |
| Depression | 6,732 (4.4%) | 162 (0.6%) | 6,570 (5.3%) | <0.001 |
| Diabetes | 10,469 (6.9%) | 107 (0.4%) | 10,362 (8.4%) | <0.001 |
| Poor diet | 19,420 (12.8%) | 482 (1.7%) | 18,938 (15.4%) | <0.001 |
| Hearing loss | 73,369 (48.3%) | 5,003 (17.4%) | 68,366 (55.5%) | <0.001 |
| Hypertension | 54,953 (36.2%) | 2,331 (8.1%) | 52,622 (42.7%) | <0.001 |
| Low physical activity | 52,691 (34.7%) | 2,220 (7.7%) | 50,471 (41.0%) | <0.001 |
| Sleep disturbance | 63,191 (41.6%) | 3,768 (13.1%) | 59,423 (48.3%) | <0.001 |
| Social isolation | 12,460 (8.2%) | 330 (1.1%) | 12,130 (9.9%) | <0.001 |
| Current smoker | 11,786 (7.8%) | 289 (1.0%) | 11,497 (9.3%) | <0.001 |
| Traumatic brain injury | 536 (0.4%) | 12 (<0.1%) | 524 (0.4%) | <0.001 |
| Visual impairment | 6,366 (4.2%) | 214 (0.7%) | 6,152 (5.0%) | <0.001 |
| Low vitamin D, <50 nmol/L | 74,737 (49.2%) | 4,644 (16.2%) | 70,093 (56.9%) | <0.001 |

P-values were calculated with the t-test for continuous variables and the chi-squared test for categorical variables. SD=standard deviation; APOE=apolipoprotein E.

During a median follow-up of 13.4 (IQR 12.7–14.0) years, 5,304 participants developed all-cause dementia. A dose‒response relationship between the number of risk factors and incident dementia was observed, with hazard ratios (HRs) of 1.10 (95% confidence interval [CI] 1.00-1.21), 1.25 (1.14-1.37), 1.39 (1.26-1.53), 1.48 (1.33-1.64) and 2.18 (1.96-2.42) for 2, 3, 4, 5, and ≥6 risk factors, respectively, compared with 0--1 risk factors (**Figure 1, Table S3**).

**Figure 1.**
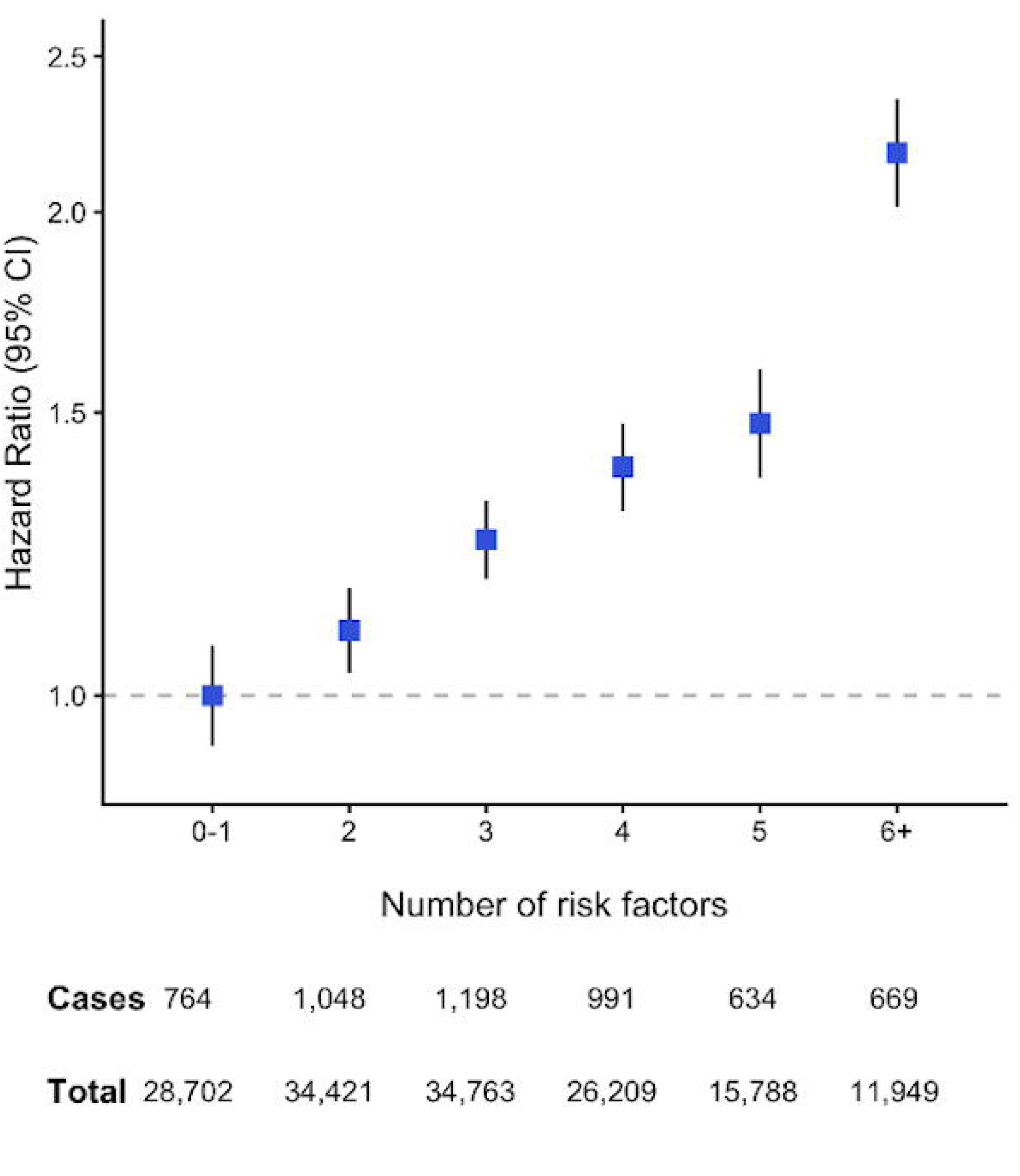
Cox proportional hazards models for the association between the number of risk factors and incident dementia (total sample) The model was adjusted for age, ethnicity, education, socioeconomic status, and APOE-ε4 carrier status. Floating absolute risks are used to estimate group-specific 95% confidence intervals for all categories. APOE=apolipoprotein E.

The LCA identified six distinct classes of risk factors among participants with ≥2 risk factors, with each class representing a unique risk factor combination (**Figure S1**). Class 1 (23.8% of the participants with ≥2 risk factors) was characterised as ‘sleep disturbance and psychosocial factors’; class 2 (22.8%) as ‘poor cardiometabolic health’; class 3 (14.1%) as ‘hearing loss and unhealthy lifestyle’; class 4 (18.7%) as ‘heavy drinking and unhealthy lifestyle’; class 5 (12.1%) as ‘vitamin D deficiency, sedentary and psychosocial factors’; and class 6 (8.5%) as ‘multiple risk factors’ (see **Table S4** for class characterisation by each risk factor). There was high similarity in the characteristics of the classes identified in the training samples versus those observed in the test sample under a six-class solution (**Table S4**). All baseline characteristics, except age and APOE-e4 carrier status, varied by class (**Table S5**). Baseline characteristics were similar between training and test datasets (**Table S6**).

The strongest association with dementia risk was observed for the ‘multiple risk factors’ class (HR=2.38, 95% CI 2.11-2.68), a class that was characterised not by specific risk factors but rather by a comparatively high number of risk factors, with a median of six (**Figure 2, Table S7**). Among the other classes, ‘sleep and psychosocial factors’ (HR=1.34, 95% CI 1.20-1.49), ‘poor cardiometabolic health’ (HR=1.32, 95% CI 1.19-1.46), ‘hearing loss and unhealthy lifestyle’ (HR=1.37, 95% CI 1.22-1.55), ‘heavy drinking and unhealthy lifestyle’ (HR=1.20, 95% CI 1.07-1.35), and ‘vitamin D deficiency, sedentary behaviour and psychosocial factors’ (HR=1.18, 95% CI 1.03-1.35) were associated with an increased risk of dementia. The direction and magnitude of associations were largely consistent across datasets. However, some did not reach statistical significance in the smaller test sample, likely reflecting reduced statistical power (n=30,364, **Figure S3**).

**Figure 2.**
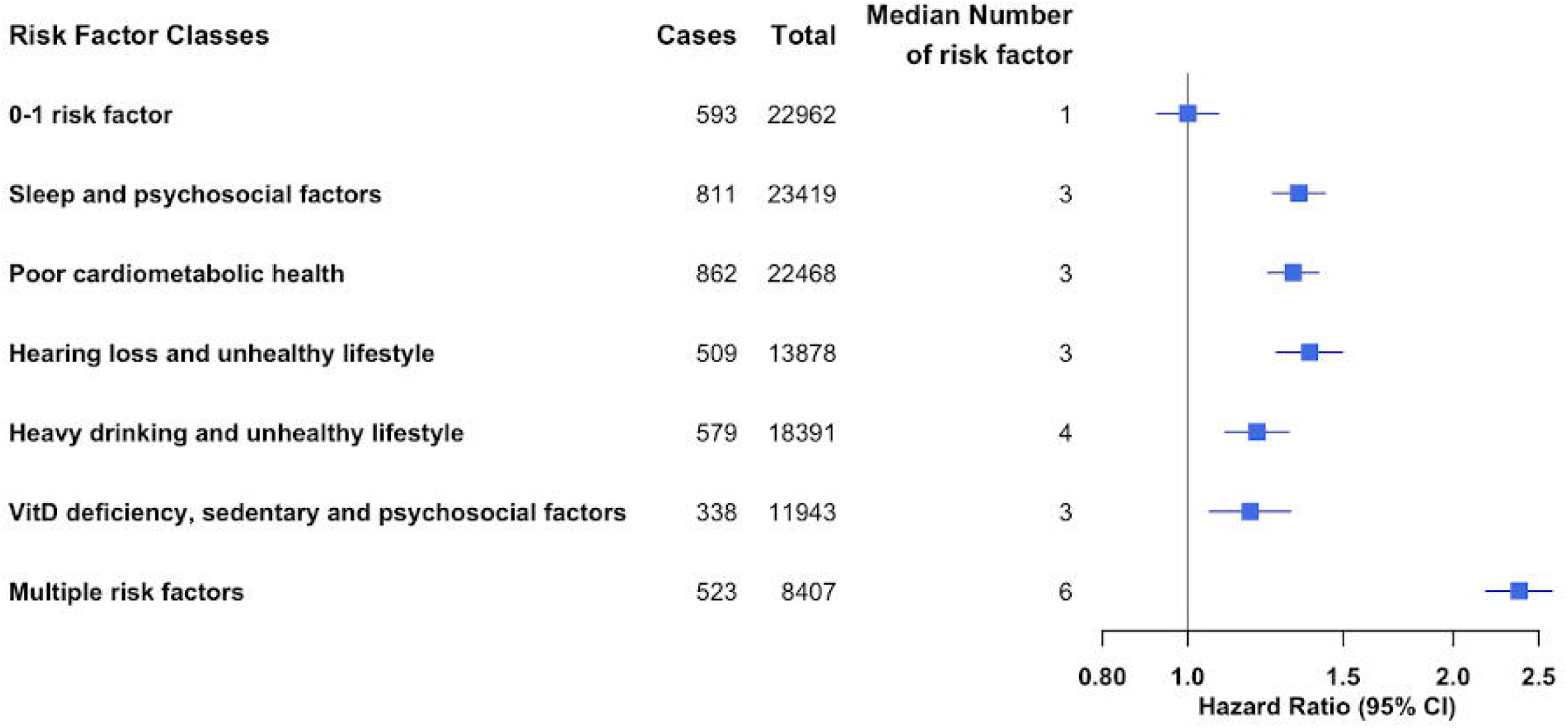
Cox proportional hazards models for the association between risk factor classes and incident dementia (training dataset) The model was adjusted for age, sex, ethnicity, education, socioeconomic status, and APOE-ε4 carrier status. Floating absolute risks are used to estimate group-specific 95% confidence intervals for all categories. Each class was characterised by risk factor patterns identified by latent class analysis. Training dataset (n=121,468) was used for this analysis. APOE=apolipoprotein E; VitD=vitamin D.

In the analysis stratified by follow-up period, the findings were largely consistent up to 10 years and after 10 years of follow-up (**Figure 3, Table S8**). The exception was for the ‘vitamin D deficiency, sedentary and psychosocial factors’ class, where the association with dementia risk up to 10 years was similar to that in the main analysis (HR=1.35, 95% CI 1.10-1.65) but substantially attenuated after 10 years (HR=1.05, 95% CI 0.88-1.26).

**Figure 3.**
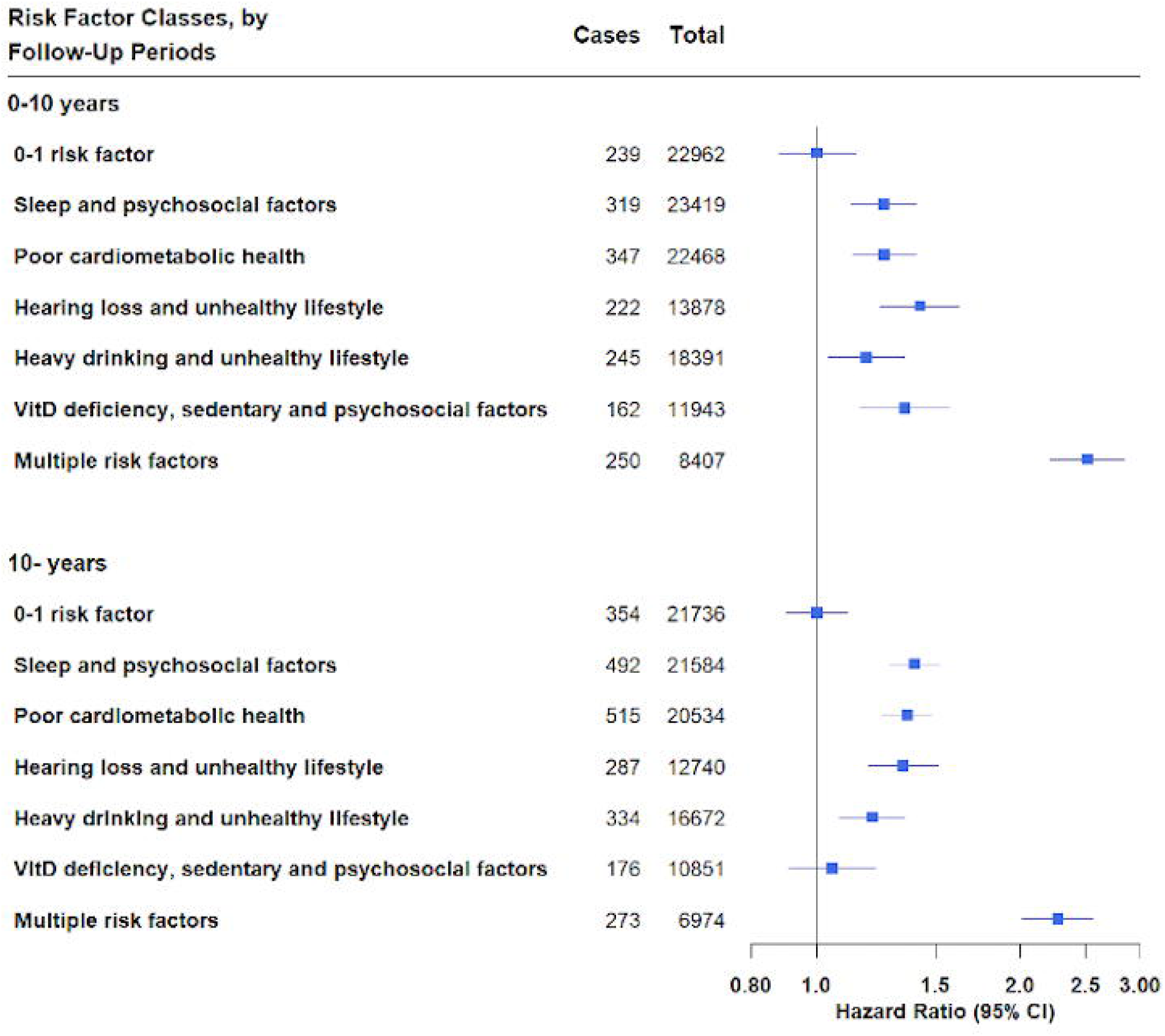
Cox proportional hazards models for the association between risk factor classes and incident dementia by different follow-up periods (training dataset) All models adjusted for age, sex, ethnicity, education, socioeconomic status, and APOE-ε4 carrier status. Floating absolute risks are used to estimate group-specific 95% confidence intervals for all categories. Each class was characterised by risk factor patterns identified by latent class analysis. Training dataset (n=121,468) was used for this analysis. APOE=apolipoprotein E; VitD=vitamin D.

APOE-ε4 carrier status significantly modified the associations between risk factor classes and dementia (**Figure 4, Table S8**). The direction of associations remained similar in the APOE-ε4 subgroup, but the relationships between each risk factor class and incident dementia were stronger among APOE-ε4 noncarriers. For example, for the association between ‘multiple risk factors’ and dementia, the HRs were 3.55 (95% CI 2.93-4.22) and 1.59 (95% 1.33-1.90) among APOE-ε4 noncarriers and APOE-ε4 carriers, respectively. The associations with dementia were also statistically nonsignificant among APOE-ε4 carriers for ‘heavy drinking and unhealthy lifestyle’ (HR=1.04, 95% CI 0.89-1.23) and ‘vitamin D deficiency, sedentary and psychosocial factors’ (HR=1.15, 95% CI 0.96-1.38). Significant interactions between APOE-ε4 carrier status and risk factor classes were observed for ‘poor cardiometabolic health’ (p = 0.0020), ‘heavy drinking and unhealthy lifestyle’ (p = 0.010), and ‘multiple risk factors’ (p < 0.001). Interactions were not statistically significant for ‘sleep and psychosocial factors’ (p = 0.13), ‘hearing loss and unhealthy lifestyle’ (p = 0.058), and ‘vitamin D deficiency, sedentary and psychosocial factors’ (p = 0.54). No significant interactions were detected between sex and any of the risk factor classes with respect to dementia risk (**Table S9**, p for interaction >0.05).

**Figure 4.**
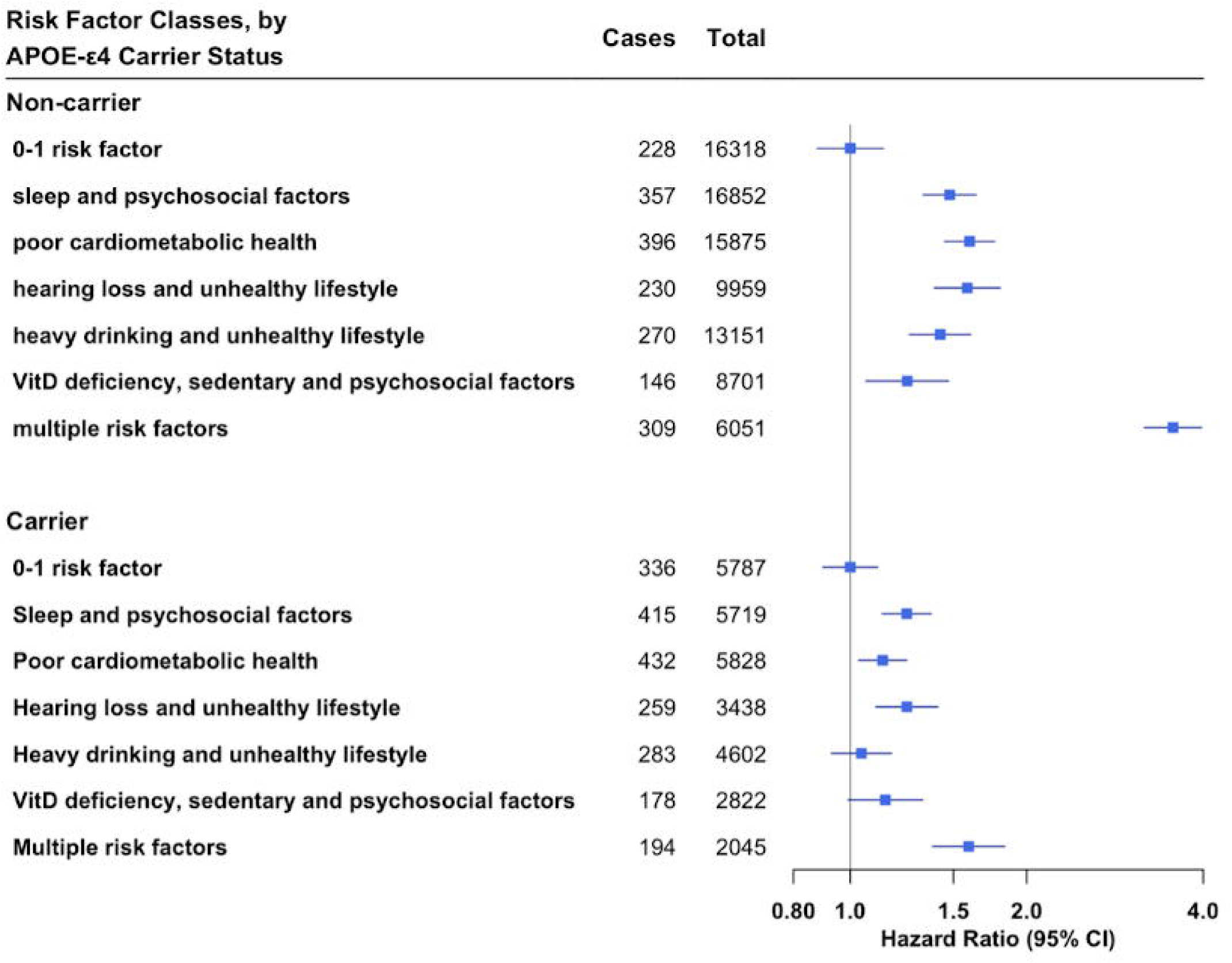
Cox proportional hazards models for the association between risk factor classes and incident dementia by APOE-ε4 carrier status (training dataset) All models adjusted for age, sex, ethnicity, education, and socioeconomic status. Floating absolute risks are used to estimate group-specific 95% confidence intervals for all categories. Each class was characterised by risk factor patterns identified by latent class analysis. P-values for interaction between APOE-ε4 carrier status and risk factor classes: Sleep and psychosocial factors p=0.13; Poor cardiometabolic health p=0.0020; Hearing loss and unhealthy lifestyle p=0.058; Heavy drinking and unhealthy lifestyle p=0.010; Vitamin D deficiency, sedentary and psychosocial factors p=0.54; Multiple risk factors p<0.001. Training dataset (n=121,468) was used for this analysis. APOE=apolipoprotein E; VitD=vitamin

The findings were robust across multiple sensitivity analyses. Results were virtually identical when attained age was used as the time scale (**Table S10**). When participants with prevalent CVD at baseline were excluded, associations were modestly attenuated but remained statistically significant for all classes, with the largest attenuation observed for the ‘multiple risk factors’ class (HR = 2.09, 95% CI 1.82–2.40) and ‘poor cardiometabolic health’ class (HR = 1.22, 95% CI 1.09–1.37), suggesting partial mediation through cardiovascular pathways (**Table S11**). When the reference group was restricted to participants with zero risk factors, hazard ratios were larger across all classes, but the relative ranking and pattern of associations remained unchanged (**Table S12**).”

## Discussion

In a population-based cohort of approximately 150,000 60–69-year-olds, we identified six distinct classes from 14 preselected lifestyle, behavioural and health-related risk factors for dementia. Compared with those with fewer than two risk factors, participants with ‘multiple risk factors’ had almost two and a half times greater risk of developing dementia over 15 years of follow-up. Distinct patterns of risk factors, including ‘sleep and psychosocial factors’, ‘poor cardiometabolic health’, ‘hearing loss and unhealthy lifestyle’, ‘heavy drinking and unhealthy lifestyle’, ‘vitamin D deficiency, sedentary behaviour and psychosocial factors’, were associated with a 20–30% increased risk of dementia. The associations remained similar, albeit slightly attenuated, when restricted to dementia cases that occurred 10 years after baseline, whereas the associations were strongest among individuals without a genetic predisposition to dementia on the basis of APOE–ε4 carrier status. Exclusion of participants with prevalent CVD resulted in modest attenuation of associations across classes, suggesting that cardiovascular pathways may partially contribute to the observed associations. Consistent with previous studies, we observed a dose‒response association between the number of risk factors and dementia incidence.^3^ However, this approach treats all factors as contributing equally and does not account for potentially distinct patterns. The LCA complements this by revealing that specific combinations of co-occurring risk factors are differentially associated with dementia risk, providing additional insight beyond a simple count. We identified a class in the cluster analysis for which the prevalence of each risk factor, except heavy drinking, was higher than expected. The participants in this ‘multiple risk factor’ class had six risk factors on average, and the class was associated with the strongest risk of dementia compared with the other five classes. This suggests that there is a subset of individuals who are at high risk of dementia because they are exposed to many risk factors rather than any particular combination. However, only 5.5% of the sample was in the ‘multiple risk factors’ class, and we identified distinct patterns of risk factors that were differentially associated with dementia risk among the remaining population. Notably, the pattern of risk factors appeared to matter beyond their number: the ‘heavy drinking and unhealthy lifestyle’ class (median of four risk factors) showed a lower risk than the ‘poor cardiometabolic health’ class (median of three), suggesting that the specific combination of co-occurring factors, rather than their count alone, may play an important role in determining dementia risk. Our findings complement and extend previous cluster-based research. Xiong and colleagues, also using UK Biobank data, reported three sex-specific profiles (cardiometabolic, substance use, and low risk), with dementia risk being highest in the cardiometabolic group. ^8^ Our study advances this literature by restricting LCA to individuals with two or more risk factors and comparing them with a reference group of individuals with zero or one risk factor. This yielded clinically relevant and diverse clusters, highlighting heterogeneous pathways associated with dementia risk. The class with the next strongest association was driven by hearing loss and consisted of a higher prevalence of certain unhealthy lifestyle factors, including poor diet, physical inactivity and smoking, as well as depression and social isolation. The 2024 Lancet Commission on dementia prevention ascribed the highest population attributable fraction to hearing, estimating that 7% of dementia cases could be caused by hearing impairment.^2^ Lifestyle factors have been linked to hearing loss^20^, and it has been hypothesised that hearing impairment could increase the likelihood of depression and social isolation, which in turn increases dementia risk.^21^ A class characterised by sleep disturbance and psychosocial problems presented the third strongest association with dementia. These factors often cooccur, with sleep quality bidirectionally related to poor psychosocial health,^22, 23^ which could synergistically increase the risk of dementia. Whether sleep is a risk factor for dementia or represents an early-stage marker of prodromal dementia is unclear.^2^ We found that the association between ‘sleep and psychosocial factors’ was stronger when the study was restricted to dementia cases that occurred more than 10 years later. However, if sleep was a marker for dementia, we would expect the relationship to attenuate over longer periods of follow-up.

A class driven by hypertension, diabetes, and obesity was associated with a 32% increased risk of dementia, despite having a lower than expected prevalence of all other risk factors, including those known to cause poor cardiometabolic health, such as heavy alcohol consumption, smoking, physical inactivity and poor diet. This class could consist of individuals who have made healthy lifestyle changes to better manage health conditions and who, while the risk of dementia remains, is still substantially lower than the ‘multiple risk factors’ class, which is characterised by individuals with both poor cardiometabolic health and unhealthy lifestyle behaviours. This finding is consistent with a recent study in the UK Biobank, which revealed that a healthy lifestyle substantially attenuated the risk of dementia among individuals with multiple cardiometabolic conditions.^24^ In contrast, the ‘heavy drinking and unhealthy lifestyle’ class, which was associated with a 20% increased risk of dementia, had a higher than expected prevalence of heavy alcohol intake, smoking and poor diet but a low prevalence of cardiometabolic conditions. A previous UK Biobank study also revealed that unhealthy lifestyles were associated with dementia risk independent of preexisting cardiometabolic disease.^24^ The remaining class associated with dementia risk was characterised primarily by low vitamin D, sedentary (physical inactivity, social isolation) and unhealthy lifestyle behaviours (smoking, poor diet). Low vitamin D might be a consequence of these other behavioural factors, i.e., through less sunlight exposure, and while vitamin D has been linked with dementia risk, evidence generally suggests that vitamin D is a marker, rather than cause, of poor health.^25, 26^ This is supported by our finding that the association between the vitamin D-driven class and dementia was substantially attenuated, becoming statistically nonsignificant, when the study was restricted to dementia cases that developed after 10 years of follow-up.

A strength of this study is the use of a population-based cohort with detailed data collection that captures a wide range of dementia risk factors. The large sample size and high number of incident dementia cases provided the statistical power necessary to identify risk factor classes robustly and investigate the associations with dementia over 15 years of follow-up.

This study also has several limitations. First, the UK Biobank is predominantly white and consists of individuals with healthier profiles and higher socioeconomic status than the general population does, which might limit the identification of risk factor classes that are more prevalent in other populations. The 14 risk factors were selected a priori from a comprehensive umbrella review, and while the dose–response association observed in this cohort supports their relevance, the identified co-occurrence patterns may differ in other populations; validation in independent and more diverse cohorts represents an important next step. Second, reliance on self-reported data for most risk factors introduces the possibility of recall error, potentially biasing effect estimates toward the null. Third, dementia was identified via hospital inpatient and death registry records, resulting in the under ascertainment of cases diagnosed in a primary care setting. However, previous research has revealed high positive predictive value of dementia in the UK Biobank as well as consistency in risk factor–dementia associations compared with the inclusion of primary care data.^13, 27^ Associations with key dementia subtypes, such as Alzheimer’s disease and vascular dementia, were not considered because of the lower accuracy of the available medical record data in identifying dementia subtypes.^13^ This is an important limitation, particularly given that APOE-ε4 is more strongly associated with Alzheimer’s disease than vascular dementia, and the observed effect modification by APOE-ε4 status should be interpreted accordingly. Future studies with validated subtype diagnoses should investigate whether risk factor patterns are differentially associated with specific dementia subtypes. Fourth, we restricted our sample to a complete case sample, resulting in 30% of participants being excluded because of missing data. Multiple imputation was not employed because imputed values could introduce artificial patterns into the latent class structure; this approach ensures that the identified patterns reflect genuine co-occurrence in the observed data. However, these exclusions may affect the generalisability of the identified risk factor patterns, and future studies using different cohorts should examine the robustness of these findings. While the core defining features of each class were consistently replicated across training and test datasets, LCA is a probabilistic approach and minor variation in secondary features across samples is expected. Fifth, the restriction to participants aged 60–69 years excludes midlife risk factor patterns that are known to influence dementia risk decades later. Future studies should examine whether similar or distinct risk factor patterns emerge in younger populations with longer follow-up. Sixth, owing to the observational design, residual confounding and noncausal explanations for the current findings remain. The underlying mechanisms linking risk factor patterns to dementia are likely multifactorial, and further research is needed to disentangle these mechanisms.

## Conclusion

In the present study, we identified distinct patterns of cooccurring risk factors that were differentially associated with dementia risk, with the specific combination of factors appearing to matter beyond their number alone. These findings suggest that accounting for co-occurring risk factor patterns might inform risk stratification and the design of future dementia prevention strategies. Investigating whether similar patterns are identified in other more diverse populations and understanding the potential pathways underlying these associations represent key next steps for informing personalised, multidomain dementia prevention interventions.

## Data Availability

UK Biobank is an open access resource. Bona fide researchers can apply to use the UK Biobank dataset by registering and applying at http://ukbiobank.ac.uk/register-apply/. All results presented in this manuscript, including the code used to generate them, will be returned to UK Biobank within 6 months of publication at which point they are made available for researchers to request (subject to UK Biobank approval).

## Consent statement

UK Biobank received ethical approval from the National Health Service North West Centre for Research Ethics Committee (Ref: 11/NW/0382). All human subjects provided informed consent via electronic signature through the touchscreen, which was conducted in accordance with the principles of the Declaration of Helsinki.

## Authorship contribution statement

CMC, EK and TJL conceived the work. TJL acquired the data. YU analysed the data and drafted the paper. All authors made substantial contributions to the design and interpretation of the work, reviewed the paper critically for important intellectual content, gave final approval for publication, and agree to be accountable for all aspects of the work in ensuring that questions related to the accuracy or integrity of any part of the work are appropriately investigated and resolved.

## Acknowledgements

This research has been conducted using the UK Biobank Resource under Application Number 33592. We are grateful to the participants for dedicating a substantial amount of time to take part in the UK Biobank study as well as the staff who make it possible.

## References

1. World Health Organization. Dementia 2025. Available from: https://www.who.int/news-room/fact-sheets/detail/dementia.

2. Livingston G, Huntley J, Liu KY, Costafreda SG, Selbaek G, Alladi S, et al. Dementia prevention, intervention, and care: 2024 report of the Lancet standing Commission. Lancet. 2024.

3. Peters R, Booth A, Rockwood K, Peters J, D’Este C, Anstey KJ. Combining modifiable risk factors and risk of dementia: a systematic review and meta-analysis. BMJ Open. 2019;9(1):e022846.

4. Dingle SE, Bowe SJ, Bujtor M, Milte CM, Daly RM, Byles J, et al. Data-driven lifestyle patterns and risk of dementia in older Australian women. Alzheimers Dement. 2024;20(2):798–808.

5. Paolillo EW, Saloner R, VandeBunte A, Lee S, Bennett DA, Casaletto KB. Multimodal lifestyle engagement patterns support cognitive stability beyond neuropathological burden. Alzheimers Res Ther. 2023;15(1):221.

6. Moored KD, Bandeen-Roche K, Snitz BE, DeKosky ST, Williamson JD, Fitzpatrick AL, et al. Risk of Dementia Differs Across Lifestyle Engagement Subgroups: A Latent Class and Time-to-Event Analysis in Community-Dwelling Older Adults. J Gerontol B Psychol Sci Soc Sci. 2022;77(5):872–84.

7. Kontari P, Fife-Schaw C, Smith K. Clustering of Cardiometabolic Risk Factors and Dementia Incidence in Older Adults: A Cross-Country Comparison in England, the United States, and China. J Gerontol A Biol Sci Med Sci. 2023;78(6):1035–44.

8. Xiong LY, Wood Alexander M, Wong YY, Wu CY, Ruthirakuhan M, Edwards JD, et al. Latent profiles of modifiable dementia risk factors in later midlife: relationships with incident dementia, cognition, and neuroimaging outcomes. Mol Psychiatry. 2025;30(2):450–60.

9. Coley N, Giulioli C, Aisen PS, Vellas B, Andrieu S. Randomised controlled trials for the prevention of cognitive decline or dementia: A systematic review. Ageing Res Rev. 2022;82:101777.

10. Yaffe K, Vittinghoff E, Dublin S, Peltz CB, Fleckenstein LE, Rosenberg DE, et al. Effect of Personalized Risk-Reduction Strategies on Cognition and Dementia Risk Profile Among Older Adults: The SMARRT Randomized Clinical Trial. JAMA Intern Med. 2024;184(1):54–62.

11. Sudlow C, Gallacher J, Allen N, Beral V, Burton P, Danesh J, et al. UK biobank: an open access resource for identifying the causes of a wide range of complex diseases of middle and old age. PLoS Med. 2015;12(3):e1001779.

12. Jones A, Ali MU, Kenny M, Mayhew A, Mokashi V, He H, et al. Potentially Modifiable Risk Factors for Dementia and Mild Cognitive Impairment: An Umbrella Review and Meta-Analysis. Dement Geriatr Cogn Disord. 2024;53(2):91–106.

13. Wilkinson T, Schnier C, Bush K, Rannikmae K, Henshall DE, Lerpiniere C, et al. Identifying dementia outcomes in UK Biobank: a validation study of primary care, hospital admissions and mortality data. Eur J Epidemiol. 2019;34(6):557–65.

14. Calvin CM, Conroy MC, Moore SF, Kuzma E, Littlejohns TJ. Association of Multimorbidity, Disease Clusters, and Modification by Genetic Factors With Risk of Dementia. JAMA Netw Open. 2022;5(9):e2232124.

15. Townsend P, Phillimore P, Beattie A. Health and deprivation: inequality and the North: Routledge; 1988.

16. Bycroft C, Freeman C, Petkova D, Band G, Elliott LT, Sharp K, et al. The UK Biobank resource with deep phenotyping and genomic data. Nature. 2018;562(7726):203–9.

17. Qureshi D, Collister J, Allen NE, Kuzma E, Littlejohns T. Association between metabolic syndrome and risk of incident dementia in UK Biobank. Alzheimers Dement. 2024;20(1):447–58.

18. Easton DF, Peto J, Babiker AG. Floating absolute risk: an alternative to relative risk in survival and case-control analysis avoiding an arbitrary reference group. Stat Med. 1991;10(7):1025–35.

19. Linzer DA, Lewis JB. poLCA: An R Package for Polytomous Variable Latent Class Analysis. Journal of Statistical Software. 2011;42(10):1–29.

20. Tsimpida D, Kontopantelis E, Ashcroft D, Panagioti M. Socioeconomic and lifestyle factors associated with hearing loss in older adults: a cross-sectional study of the English Longitudinal Study of Ageing (ELSA). BMJ Open. 2019;9(9):e031030.

21. Shukla A, Harper M, Pedersen E, Goman A, Suen JJ, Price C, et al. Hearing Loss, Loneliness, and Social Isolation: A Systematic Review. Otolaryngol Head Neck Surg. 2020;162(5):622–33.

22. Yasugaki S, Okamura H, Kaneko A, Hayashi Y. Bidirectional relationship between sleep and depression. Neurosci Res. 2025;211:57–64.

23. Dworschak C, Mäder T, Rühlmann C, Maercker A, Kleim B. Examining bi-directional links between loneliness, social connectedness and sleep from a trait and state perspective. Sci Rep. 2024;14(1):17300.

24. Xiong S, Hou N, Tang F, Li J, Deng H. Association of cardiometabolic multimorbidity and adherence to a healthy lifestyle with incident dementia: a large prospective cohort study. Diabetol Metab Syndr. 2023;15(1):208.

25. Zhang XX, Wang HR, Meng W, Hu YZ, Sun HM, Feng YX, et al. Association of Vitamin D Levels with Risk of Cognitive Impairment and Dementia: A Systematic Review and Meta-Analysis of Prospective Studies. J Alzheimers Dis. 2024;98(2):373–85.

26. Autier P, Boniol M, Pizot C, Mullie P. Vitamin D status and ill health: a systematic review. Lancet Diabetes Endocrinol. 2014;2(1):76–89.

27. Clifton L, Liu X, Collister JA, Littlejohns TJ, Allen N, Hunter DJ. Assessing the importance of primary care diagnoses in the UK Biobank. Eur J Epidemiol. 2024;39(2):219–29.

