## Supplementary Material for "Patterns of modifiable lifestyle, behavioural and health-related risk factors and their associations with incident dementia in UK Biobank"

### Table S1. Dementia and risk factor definitions and UK Biobank data field ID

| Variables | How measured in UK Biobank measurement and variable definition | UK Biobank data field |
|---|---|---|
| Alcohol consumption | Touchscreen questionnaire at baseline. Total alcohol consumption per week was calculated based on the frequency and quantity of consumption. Respondents were categorized as non-drinkers (0), those drinking less than 21 units per week (1), and those consuming more than 21 units per week (2).<br>*Glasses were converted to UK units as follows: red or white wine = 1.5 units; fortified wine=1 unit; pint = 2 units; spirits = 1 unit; other (e.g. alcopops) =1.5 units. | 20117, 1558, 1568,1578, 1588, 1598,1608, 5364, 4407,4418, 4429, 4440,4451, 4462 |
| Obesity | Physical measurements. Body Mass Index (BMI) was constructed based on height and weight, which were measured during the initial Assessment Centre visit. Obesity was defined as BMI≥ 30 kg/m$^2$. | 21001 |
| Depression | Nurse-led verbal interview at baseline. Current or a history of depression was ascertained based on self-report. | 20002 |
| Diabetes | Nurse-led verbal interview at baseline. Current or history of diabetes was ascertained based on self-report. | 20002 |
| Diet (poor dietary pattern) | Touchscreen questionnaire at baseline.<br>Less than two of the following five food groups:<br>1. Fruits: ≥ 3 servings/day<br>2. Vegetables: ≥ 4 servings/day<br>3. Fish: ≥2 servings/week<br>4. Processed meats: ≤ 2 servings/week<br>5. Unprocessed red meats: ≤ 2 servings/week | 1309, 1319, 1289, 1299, 1329, 1339, 1349, 1369, 1379, 1389, 1438, 1448, 1458, 1468 |
| Hearing loss | Touchscreen questionnaire at baseline. Self-reported problems with hearing, including in a noisy environment; use of hearing aid. | 2247,2257, 3393 |
| Hypertension | Nurse-led verbal interview at baseline. Current or a history of hypertension was ascertained based on self-report. | 20002 |
| Physical inactivity | Touchscreen questionnaire at baseline. <150 minutes moderate activity per week OR < 75 minutes vigorous activity per week OR equivalent combination. | 884, 894, 904, 914 |
| Sleep disturbance | Touchscreen questionnaire at baseline. Based on the questions of sleep disturbance (0: never/rarely/sometimes, 1: usually) and sleep duration (0: 7-9 hours, 1: <7 or >9 hours), a binary variable was created, with one indicating either usual sleep disturbance or suboptimal sleep duration, and zero representing no significant sleep issues. | 1160, 1200 |
| Social isolation | Touchscreen questionnaire at baseline. The social isolation scale included three questions: (1) 'How many people live in your household?' (one point for living alone); (2) 'How often do you visit or receive visits from friends or family?' (one point for less than monthly visits); and (3) 'Which social activities do you attend weekly?' (one point for no weekly participation). Scores range from zero to three, with two or more points indicating social isolation. | 709, 1031,6160 |
| Smoking | Touchscreen questionnaire at baseline. Smoking status was categorized as non-current smoker (former or never smoker) and current smoker. | 20116 |
| Traumatic Brain Injury | A history of traumatic brain injury (TBI) was determined through hospital inpatient data using ICD-9 (850-854, 800-804) and ICD-10 codes (S020-S029, S060-S071, S078-S079, S097-S099, T04, T06). | 2000 |
| Visual impairment | Nurse-led verbal interview at baseline. Current or a history of visual impairment was ascertained based on self-report current or history of cataract, diabetic eye disease, glaucoma, and macular degeneration. | 20002 |
| Low Vitamin D | Blood assays. Vitamin D levels are categorized as normal (≥ 50 nmol/L) or insufficient/deficient (<50 nmol/L). | 30890 |

**Table S2. International Classification of Disease codes used to define all-cause dementia**

| ICD-9/10 code | Dementia type |
|---|---|
| 290.2 | Senile dementia, depressed or paranoid type |
| 290.3 | Senile dementia with acute confusional state |
| 290.4 | Arteriosclerotic dementia |
| 291.2 | Other alcoholic dementia |
| 294.1 | Dementia in other conditions classified elsewhere |
| 331.0 | Alzheimer's disease |
| 331.1 | Pick's disease |
| 331.2 | Senile degeneration of brain |
| 331.5 | Creutzfeldt-Jakob disease |
| A81.0 | Sporadic Creutzfeldt-Jakob disease |
| F00 | Dementia in Alzheimer's disease |
| F00.0 | Dementia in Alzheimer's disease with early onset |
| F00.1 | Dementia in Alzheimer's disease with late onset |
| F00.2 | Dementia in Alzheimer's disease, atypical or mixed type |
| F00.9 | Dementia in Alzheimer's disease, unspecified |
| F01 | Vascular dementia |
| F01.0 | Vascular dementia of acute onset |
| F01.1 | Multi-infarct dementia |
| F01.2 | Subcortical vascular dementia |
| F01.3 | Mixed cortical and sub-cortical vascular dementia |
| F01.8 | Other vascular dementia |
| F01.9 | Vascular dementia, unspecified |
| F02 | Dementia in other diseases classified elsewhere |
| F02.0 | Dementia in Picks disease |
| F02.1 | Dementia in Creutzfeldt-Jacob disease |
| F02.2 | Dementia in Huntington's disease |
| F02.3 | Dementia in Parkinson's disease |
| F02.4 | Dementia in HIV disease |
| F02.8 | Dementia in other specified diseases classified elsewhere |
| F03 | Unspecified dementia |
| F05.1 | Delirium superimposed on dementia |
| F10.6 | Mental and behavioural disorders due to use of alcohol - amnesic syndrome |
| G30 | Alzheimer's disease |
| G30.0 | Alzheimer's disease with early onset |
| G30.1 | Alzheimer's disease with late onset |
| G30.8 | Other Alzheimer's disease |
| G30.9 | Alzheimer's disease unspecified |
| G31.0 | Circumscribed brain atrophy |
| G31.1 | Senile degeneration of brain |
| G31.8 | Other specified degenerative diseases of nervous system |
| I67.3 | Binswanger's disease |

**Figure S1. Model fit statistics for class solutions (training dataset)**

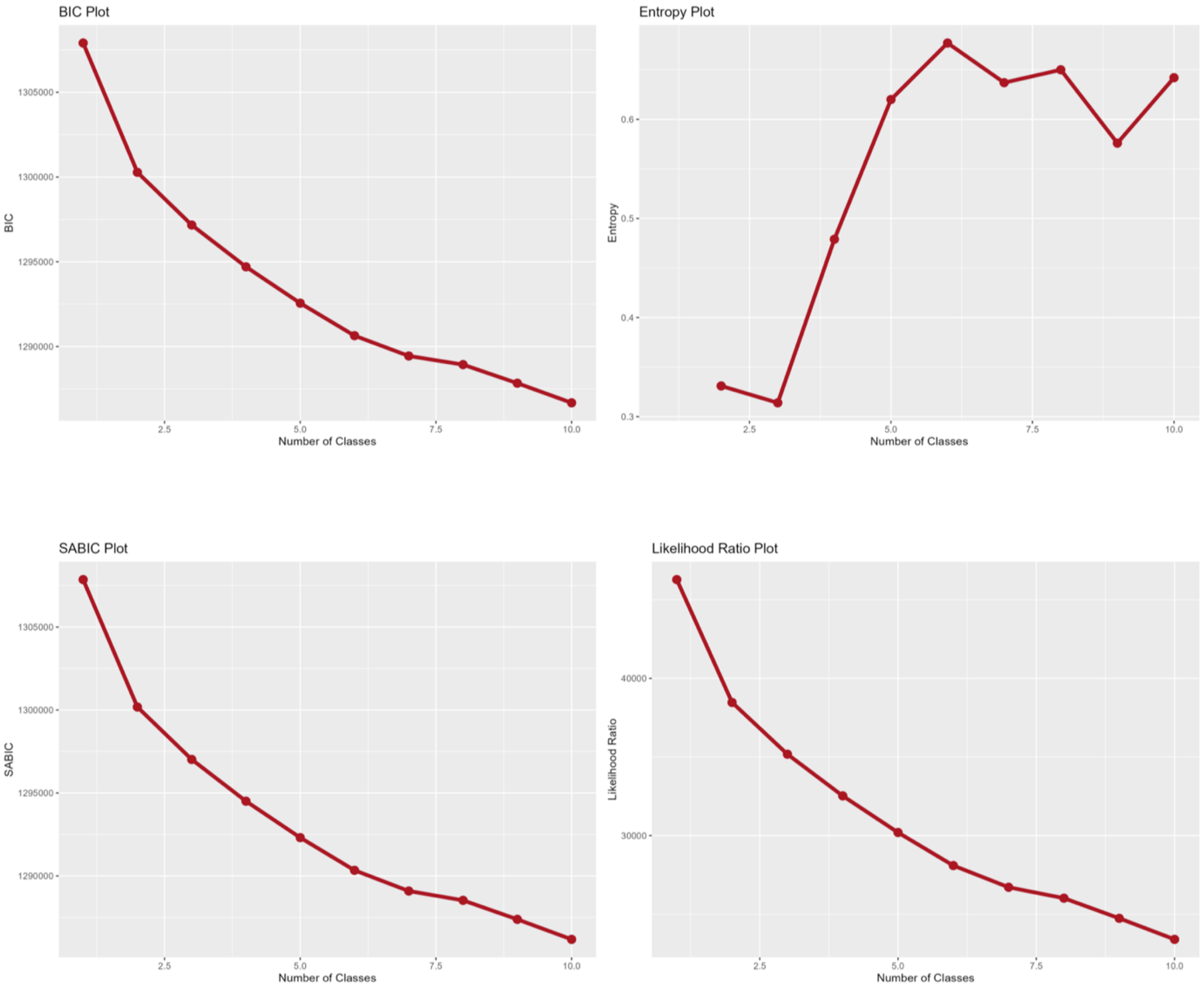


Bayesian information criteria (BIC), Sample size adjusted Bayesian information criteria (SABIC), entropy, and likelihood ratio statistics were calculated based on multiple latent class analysis models, ranging from one to ten class solutions.

Best model: 6-class model

Rationale: This solution had relatively low BIC, SABIC, and likelihood ratio statistics. Adding extra classes provided little gain in class separation (entropy) and reduced the clarity of clinical interpretation.

Training dataset (n=98,506) was used for this analysis.

**Figure S2. Cohort flow diagram**

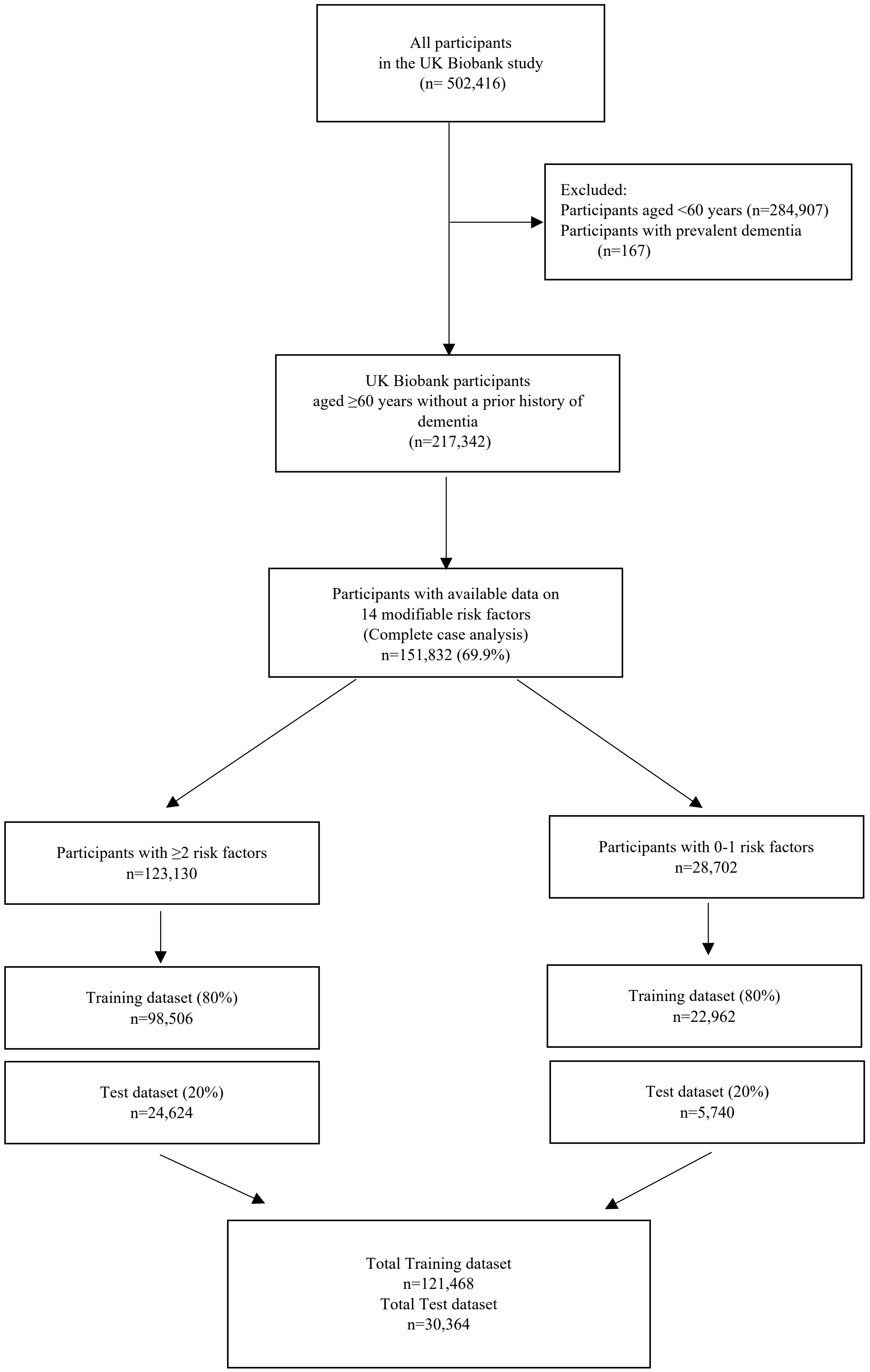

**Table S3. Cox proportional hazards models for the association between the number of risk factors and incident dementia (sequential adjustment, training dataset)**

| | Age and Sex Adjusted Model | | | Fully Adjusted Model | | |
|---|---|---|---|---|---|---|
| **Variable** | **HR** | **95% CI** | **p-value** | **HR** | **95% CI** | **p-value** |
| **Age** | 1.22 | 1.21, 1.23 | <0.0001 | 1.22 | 1.21, 1.23 | <0.0001 |
| **Sex** | | | | | | |
| **Female** | — | — | | — | — | |
| **Male** | 1.15 | 1.09, 1.22 | <0.0001 | 1.16 | 1.10, 1.23 | <0.0001 |
| **Number of risk factors** | | | | | | |
| **0-1 risk factors** | — | — | | — | — | |
| **2 risk factors** | 1.11 | 1.02, 1.22 | 0.022 | 1.10 | 1.00, 1.21 | 0.050 |
| **3 risk factors** | 1.28 | 1.17, 1.40 | <0.0001 | 1.25 | 1.14, 1.37 | <0.0001 |
| **4 risk factors** | 1.43 | 1.30, 1.57 | <0.0001 | 1.39 | 1.26, 1.53 | <0.0001 |
| **5 risk factors** | 1.56 | 1.40, 1.73 | <0.0001 | 1.48 | 1.33, 1.64 | <0.0001 |
| **6 or more risk factors** | 2.34 | 2.11, 2.60 | <0.0001 | 2.18 | 1.96, 2.42 | <0.0001 |
| **Ethnicity** | | | | | | |
| **Non-White** | | | | — | — | |
| **White** | | | | 0.85 | 0.72, 1.00 | 0.048 |
| **Missing** | | | | 0.62 | 0.34, 1.15 | 0.13 |
| **Education** | | | | | | |
| **Primary** | | | | — | — | |
| **Secondary** | | | | 0.80 | 0.74, 0.85 | <0.0001 |
| **Post-secondary non-tertiary** | | | | 0.82 | 0.75, 0.89 | <0.0001 |
| **Tertiary** | | | | 0.71 | 0.65, 0.76 | <0.0001 |
| **Missing** | | | | 1.18 | 0.94, 1.50 | 0.16 |
| **Socioeconomic status, quintile** | | | | | | |
| **Least deprived** | | | | — | — | |
| **Second least deprived** | | | | 1.01 | 0.92, 1.10 | 0.86 |
| **Middle** | | | | 1.02 | 0.93, 1.11 | 0.64 |
| **Second most deprived** | | | | 1.10 | 1.01, 1.20 | 0.028 |
| **Most deprived** | | | | 1.31 | 1.20, 1.43 | <0.0001 |
| **Missing** | | | | 1.08 | 0.40, 2.88 | 0.88 |
| **APOE-ε4 carrier status** | | | | | | |
| **non-carrier** | | | | — | — | |
| **carrier** | | | | 3.36 | 3.18, 3.55 | <0.0001 |
| **Missing** | | | | 2.05 | 1.79, 2.35 | <0.0001 |

Training dataset (n=121,468) was used for this analysis. HR=hazard ratio; CI=confidence interval; APOE=apolipoprotein E.

**Table S4. Observed prevalence and observed and expected prevalence ratio within six classes**

**1. Training dataset**

| Risk Factor | Expected Prevalence (%) | Observed Prevalence (%) Class | | | | | | The Ratio of Observed and Expected Prevalence Class | | | | | |
|---|---|---|---|---|---|---|---|---|---|---|---|---|---|
| | | 1 | 2 | 3 | 4 | 5 | 6 | 1 | 2 | 3 | 4 | 5 | 6 |
| Heavy Drinking | 25.9 | 0.0 | 19.2 | 0.0 | 100.0 | 5.3 | 24.4 | 0.0 | 0.7 | 0.0 | 3.9 | 0.2 | 0.9 |
| Obesity | 28.8 | 14.9 | 36.0 | 18.7 | 14.0 | 37.8 | 84.5 | 0.5 | 1.3 | 0.7 | 0.5 | 1.3 | 2.9 |
| Depression | 5.3 | 6.5 | 2.3 | 6.4 | 3.4 | 6.4 | 11.3 | 1.2 | 0.4 | 1.2 | 0.6 | 1.2 | 2.1 |
| Diabetes | 8.4 | 2.2 | 10.4 | 3.4 | 1.5 | 4.4 | 49.5 | 0.3 | 1.2 | 0.4 | 0.2 | 0.5 | 5.9 |
| Unhealthy Diet | 15.4 | 11.4 | 4.9 | 18.0 | 21.7 | 23.7 | 24.2 | 0.7 | 0.3 | 1.2 | 1.4 | 1.5 | 1.6 |
| Hearing Loss | 55.5 | 62.9 | 42.2 | 100.0 | 63.3 | NA | 59.1 | 1.1 | 0.8 | 1.8 | 1.1 | NA | 1.1 |
| Hypertension | 42.7 | 14.6 | 100.0 | 11.6 | 24.9 | 22.1 | 89.1 | 0.3 | 2.3 | 0.3 | 0.6 | 0.5 | 2.1 |
| Social Isolation | 9.9 | 11.4 | 2.2 | 10.4 | 4.4 | 19.5 | 23.8 | 1.2 | 0.2 | 1.1 | 0.4 | 2.0 | 2.4 |
| Physical Inactivity | 41.0 | 37.7 | 23.1 | 45.5 | 33.4 | 67.7 | 69.0 | 0.9 | 0.6 | 1.1 | 0.8 | 1.7 | 1.7 |
| Traumatic Brain Injury | 0.4 | 0.4 | 0.4 | 0.5 | 0.4 | 0.6 | 0.4 | 0.8 | 0.9 | 1.1 | 1.0 | 1.4 | 1.0 |
| Sleep Disturbance | 48.3 | 100.0 | 39.6 | NA | 41.4 | 13.3 | 71.6 | 2.1 | 0.8 | NA | 0.9 | 0.3 | 1.5 |
| Current Smoking | 9.3 | 7.9 | 0.8 | 10.2 | 14.6 | 17.1 | 11.9 | 0.8 | 0.1 | 1.1 | 1.6 | 1.8 | 1.3 |
| Visual Impairment | 5.0 | 4.0 | 4.6 | 5.2 | 3.3 | 6.4 | 9.9 | 0.8 | 0.9 | 1.0 | 0.7 | 1.3 | 2.0 |
| Low Vitamin D | 56.9 | 54.0 | 38.6 | 64.9 | 46.8 | 85.1 | 83.8 | 0.9 | 0.7 | 1.1 | 0.8 | 1.5 | 1.5 |

**2. Test dataset**

| Risk Factor | Expected Prevalence (%) | Observed Prevalence (%) Class | | | | | | The Ratio of Observed and Expected Prevalence Class | | | | | |
|---|---|---|---|---|---|---|---|---|---|---|---|---|---|
| | | 1 | 2 | 3 | 4 | 5 | 6 | 1 | 2 | 3 | 4 | 5 | 6 |
| Heavy Drinking | 25.9 | 0.7 | 9.8 | NA | 100.0 | 0.7 | 26.7 | 0.0 | 0.4 | NA | 3.9 | 0.0 | 1.0 |
| Obesity | 28.8 | 16.4 | 31.1 | 18.5 | 13.7 | 36.1 | 88.7 | 0.6 | 1.1 | 0.6 | 0.5 | 1.3 | 3.1 |
| Depression | 5.3 | 7.4 | 1.9 | 6.1 | 3.4 | 4.9 | 11.3 | 1.4 | 0.3 | 1.1 | 0.6 | 0.9 | 2.1 |
| Diabetes | 8.4 | 2.2 | 10.4 | 3.3 | 1.3 | 4.5 | 45.1 | 0.3 | 1.2 | 0.4 | 0.2 | 0.5 | 5.4 |
| Unhealthy Diet | 15.4 | 13.0 | 4.7 | 19.1 | 19.7 | 22.1 | 22.2 | 0.8 | 0.3 | 1.2 | 1.3 | 1.4 | 1.4 |
| Hearing Loss | 55.5 | 61.6 | 49.9 | 100.0 | 56.7 | NA | 59.4 | 1.1 | 0.9 | 1.8 | 1.0 | NA | 1.1 |
| Hypertension | 42.7 | 5.2 | 100.0 | 10.0 | 30.5 | 22.9 | 88.0 | 0.1 | 2.3 | 0.2 | 0.7 | 0.5 | 2.1 |
| Social Isolation | 9.9 | 11.6 | 3.0 | 9.6 | 5.0 | 18.5 | 19.4 | 1.2 | 0.3 | 1.0 | 0.5 | 1.9 | 2.0 |
| Physical Inactivity | 41.0 | 36.2 | 24.5 | 46.0 | 34.7 | 65.2 | 65.5 | 0.9 | 0.6 | 1.1 | 0.8 | 1.6 | 1.6 |
| Traumatic Brain Injury | 0.4 | 0.4 | 0.5 | 0.5 | 0.4 | 0.3 | 0.5 | 0.9 | 1.2 | 1.3 | 0.9 | 0.8 | 1.2 |
| Sleep Disturbance | 48.3 | 100.0 | 47.9 | NA | 39.6 | 16.8 | 61.9 | 2.1 | 1.0 | NA | 0.8 | 0.3 | 1.3 |
| Current Smoking | 9.3 | 8.1 | 1.5 | 9.6 | 14.1 | 17.4 | 9.3 | 0.9 | 0.2 | 1.0 | 1.5 | 1.9 | 1.0 |
| Visual Impairment | 5.0 | 3.8 | 4.6 | 5.4 | 3.9 | 8.1 | 8.0 | 0.8 | 0.9 | 1.1 | 0.8 | 1.6 | 1.6 |
| Low Vitamin D | 56.9 | 51.3 | 35.4 | 65.5 | 48.2 | 85.7 | 85.8 | 0.9 | 0.6 | 1.2 | 0.8 | 1.5 | 1.5 |

Class 1: sleep and psychosocial factors; Class 2: poor cardiometabolic health; Class 3: hearing loss and unhealthy lifestyle; Class 4: heavy drinking and unhealthy lifestyle; Class 5: vitamin D deficiency, sedentary behaviour, and psychosocial factors; Class 6: multiple risk factors. Each class was identified by latent class analysis and characterised by risk factor patterns with higher observed prevalence, excluding risk factors for which their ratio of observed and expected prevalence was less than 1 (i.e., the observed prevalence was less than the expected prevalence). The analysis focused on the participants with two or more risk factors.

**Table S5. Baseline characteristics by risk factor classes (training dataset)**

| Characteristics | Overall, 121,468 | 0-1 risk factors N=22,962 | Class 1 N=23,419 | Class 2 N=22,468 | Class 3 N=13,878 | Class 4 N=18,391 | Class 5 N=11,943 | Class 6 N=8,407 |
|---|---|---|---|---|---|---|---|---|
| **Age, years** | 64.1 (2.8) | 64.0 (2.8) | 64.1 (2.9) | 64.5 (2.8) | 64.2 (2.9) | 63.9 (2.8) | 63.7 (2.8) | 64.2 (2.9) |
| **Sex** | | | | | | | | |
| **Female** | 59,728 (49.2%) | 14,188 (61.8%) | 13,897 (59.3%) | 10,696 (47.6%) | 6,538 (47.1%) | 4,160 (22.6%) | 6,832 (57.2%) | 3,417 (40.6%) |
| **Male** | 61,740 (50.8%) | 8,774 (38.2%) | 9,522 (40.7%) | 11,772 (52.4%) | 7,340 (52.9%) | 14,231 (77.4%) | 5,111 (42.8%) | 4,990 (59.4%) |
| **Ethnicity** | | | | | | | | |
| **Non-White** | 2,883 (2.4%) | 282 (1.2%) | 655 (2.8%) | 688 (3.1%) | 324 (2.3%) | 147 (0.8%) | 355 (3.0%) | 432 (5.1%) |
| **White** | 118,226 (97.3%) | 22,632 (98.6%) | 22,692 (96.9%) | 21,714 (96.6%) | 13,507 (97.3%) | 18,188 (98.9%) | 11,550 (96.7%) | 7,943 (94.5%) |
| **Missing** | 359 (0.3%) | 48 (0.2%) | 72 (0.3%) | 66 (0.3%) | 47 (0.3%) | 56 (0.3%) | 38 (0.3%) | 32 (0.4%) |
| **Education** | | | | | | | | |
| **Primary** | 28,528 (23.5%) | 4,525 (19.7%) | 5,894 (25.2%) | 5,774 (25.7%) | 3,025 (21.8%) | 3,731 (20.3%) | 2,752 (23.0%) | 2,827 (33.6%) |
| **Secondary** | 39,552 (32.6%) | 8,074 (35.2%) | 7,591 (32.4%) | 7,201 (32.1%) | 4,335 (31.2%) | 5,749 (31.3%) | 4,070 (34.1%) | 2,532 (30.1%) |
| **Post-secondary non-tertiary** | 17,653 (14.5%) | 3,163 (13.8%) | 3,320 (14.2%) | 3,531 (15.7%) | 2,010 (14.5%) | 2,726 (14.8%) | 1,674 (14.0%) | 1,229 (14.6%) |
| **Tertiary** | 34,730 (28.6%) | 7,004 (30.5%) | 6,405 (27.3%) | 5,737 (25.5%) | 4,390 (31.6%) | 6,087 (33.1%) | 3,374 (28.3%) | 1,733 (20.6%) |
| **Missing** | 1,005 (0.8%) | 196 (0.9%) | 209 (0.9%) | 225 (1.0%) | 118 (0.9%) | 98 (0.5%) | 73 (0.6%) | 86 (1.0%) |
| **Socioeconomic status, quintile** | | | | | | | | |
| **Least deprived** | 25,343 (20.9%) | 5,726 (24.9%) | 4,629 (19.8%) | 4,733 (21.1%) | 2,894 (21.4%) | 3,943 (21.4%) | 2,332 (19.5%) | 1,086 (12.9%) |
| **Second least deprived** | 24,951 (20.5%) | 5,267 (22.9%) | 4,713 (20.1%) | 4,838 (21.5%) | 2,854 (20.6%) | 3,762 (20.5%) | 2,266 (19.0%) | 1,251 (14.9%) |
| **Middle** | 24,597 (20.2%) | 4,926 (21.5%) | 4,755 (20.3%) | 4,682 (20.8%) | 2,894 (20.9%) | 3,645 (19.8%) | 2,279 (19.1%) | 1,416 (16.8%) |
| **Second most deprived** | 24,130 (19.9%) | 4,185 (18.2%) | 4,766 (20.4%) | 4,436 (19.7%) | 2,816 (20.3%) | 3,650 (19.8%) | 2,474 (20.7%) | 1,803 (21.4%) |
| **Most deprived** | 22,344 (18.4%) | 2,840 (12.4%) | 4,528 (19.3%) | 3,763 (16.7%) | 2,409 (17.4%) | 3,381 (18.4%) | 2,582 (21.6%) | 2,841 (33.8%) |
| **Missing** | 103 (0.1%) | 18 (0.1%) | 28 (0.1%) | 16 (0.1%) | 11 (0.1%) | 10 (0.1%) | 10 (0.1%) | 10 (0.1%) |
| **APOE-ε4 carrier status** | | | | | | | | |
| **non-carrier** | 86,907 (71.5%) | 16,318 (71.1%) | 16,852 (72.0%) | 15,875 (70.7%) | 9,959 (71.8%) | 13,151 (71.5%) | 8,701 (72.9%) | 6,051 (72.0%) |
| **carrier** | 30,241 (24.9%) | 5,787 (25.2%) | 5,719 (24.4%) | 5,828 (25.9%) | 3,438 (24.8%) | 4,602 (25.0%) | 2,822 (23.6%) | 2,045 (24.3%) |
| **Missing** | 4,320 (3.6%) | 857 (3.7%) | 848 (3.6%) | 765 (3.4%) | 481 (3.5%) | 638 (3.5%) | 420 (3.5%) | 311 (3.7%) |

Class 1: sleep and psychosocial factors; Class 2: poor cardiometabolic health; Class 3: hearing loss and unhealthy lifestyle; Class 4: heavy drinking and unhealthy lifestyle; Class 5: vitamin D deficiency, sedentary behaviour, and psychosocial factors; Class 6: multiple risk factors. SD=standard deviation; IQR=interquartile range.
Training dataset (n=121,468) was used for this analysis.

**Table S6. Baseline characteristics by risk factor classes (test dataset)**

| Characteristics | Overall, 30,364 | 0-1 risk factors N=5,740 | Class 1 N=5,237 | Class 2 N=5,396 | Class 3 N=3,369 | Class 4 N=5,217 | Class 5 N=2,990 | Class 6 N=2,415 |
|---|---|---|---|---|---|---|---|---|
| **Age, years** | 64.1 (2.9) | 63.9 (2.8) | 64.0 (2.9) | 64.6 (2.9) | 64.2 (2.9) | 63.9 (2.8) | 63.7 (2.8) | 64.2 (2.8) |
| **Sex** | | | | | | | | |
| **Female** | 15,029(49.5%) | 3,516 (61.3%) | 3,131 (59.8%) | 2,807 (52.0%) | 1,618 (48.0%) | 1,180 (22.6%) | 1,791 (59.9%) | 986 (40.8%) |
| **Male** | 15,335(50.5%) | 2,224 (38.7%) | 2,106 (40.2%) | 2,589 (48.0%) | 1,751 (52.0%) | 4,037 (77.4%) | 1,199 (40.1%) | 1,429 (59.2%) |
| **Ethnicity** | | | | | | | | |
| **Non-White** | 702 (2.3%) | 68 (1.2%) | 140 (2.7%) | 169 (3.1%) | 89 (2.6%) | 34 (0.7%) | 90 (3.0%) | 112 (4.6%) |
| **White** | 29,581 (97.4%) | 5,660 (98.6%) | 5,082 (97.0%) | 5,205 (96.5%) | 3,266 (96.9%) | 5,175 (99.2%) | 2,896 (96.9%) | 2,297 (95.1%) |
| **Missing** | 81 (0.3%) | 12 (0.2%) | 15 (0.3%) | 22 (0.4%) | 14 (0.4%) | 8 (0.2%) | 4 (0.1%) | 6 (0.2%) |
| **Education** | | | | | | | | |
| **Primary** | 7,214 (23.8%) | 1,148 (20.0%) | 1,383 (26.4%) | 1,460 (27.1%) | 759 (22.5%) | 1,046 (20.0%) | 686 (22.9%) | 732 (30.3%) |
| **Secondary** | 10,002 (32.9%) | 2,029 (35.3%) | 1,767 (33.7%) | 1,729 (32.0%) | 1,090 (32.4%) | 1,655 (31.7%) | 969 (32.4%) | 763 (31.6%) |
| **Post-secondary non-tertiary** | 4,353 (14.3%) | 786 (13.7%) | 684 (13.1%) | 847 (15.7%) | 462 (13.7%) | 742 (14.2%) | 448 (15.0%) | 384 (15.9%) |
| **Tertiary** | 8,566 (28.2%) | 1,735 (30.2%) | 1,363 (26.0%) | 1,308 (24.2%) | 1,040 (30.9%) | 1,751 (33.6%) | 855 (28.6%) | 514 (21.3%) |
| **Missing** | 229 (0.8%) | 42 (0.7%) | 40 (0.8%) | 52 (1.0%) | 18 (0.5%) | 23 (0.4%) | 32 (1.1%) | 22 (0.9%) |
| **Socioeconomic status, quintile** | | | | | | | | |
| **Least deprived** | 6,343 (20.9%) | 1,439 (25.1%) | 1,069 (20.4%) | 1,114 (20.6%) | 738 (21.9%) | 1,073 (20.6%) | 576 (19.3%) | 334 (13.8%) |
| **Second least deprived** | 6,348 (20.9%) | 1,322 (23.0%) | 1,039 (19.8%) | 1,190 (22.1%) | 692 (20.5%) | 1,116 (21.4%) | 588 (19.7%) | 401 (16.6%) |
| **Middle** | 6,097 (20.1%) | 1,231 (21.4%) | 1,003 (19.2%) | 1,080 (20.0%) | 677 (20.1%) | 1,065 (20.4%) | 634 (21.2%) | 407 (16.9%) |
| **Second most deprived** | 6,015 (19.8%) | 1,050 (18.3%) | 1,112 (21.2%) | 1,060 (19.6%) | 662 (19.6%) | 1,030 (19.7%) | 568 (19.0%) | 533 (22.1%) |
| **Most deprived** | 5,538 (18.2%) | 697 (12.1%) | 1,008 (19.2%) | 948 (17.6%) | 599 (17.8%) | 927 (17.8%) | 622 (20.8%) | 737 (30.5%) |
| **Missing** | 23 (0.1%) | 1 (0.0%) | 6 (0.1%) | 4 (0.1%) | 1 (0.0%) | 6 (0.1%) | 2 (0.1%) | 3 (0.1%) |
| **APOE-ε4 carrier status** | | | | | | | | |
| **non-carrier** | 21,739 (71.6%) | 4,108 (71.6%) | 3,754 (71.7%) | 3,798 (70.4%) | 2,388 (70.9%) | 3,790 (72.6%) | 2,166 (72.4%) | 1,735 (71.8%) |
| **carrier** | 7,527 (24.8%) | 1,407 (24.5%) | 1,293 (24.7%) | 1,413 (26.2%) | 840 (24.9%) | 1,260 (24.2%) | 733 (24.5%) | 581 (24.1%) |
| **Missing** | 1,098 (3.6%) | 225 (3.9%) | 190 (3.6%) | 185 (3.4%) | 141 (4.2%) | 167 (3.2%) | 91 (3.0%) | 99 (4.1%) |

Class 1: sleep and psychosocial factors; Class 2: poor cardiometabolic health; Class 3: hearing loss and unhealthy lifestyle; Class 4: heavy drinking and unhealthy lifestyle; Class 5: vitamin D deficiency, sedentary behaviour, and psychosocial factors; Class 6: multiple risk factors. SD=standard deviation; IQR=interquartile range.
Test dataset (n=30,364) was used for this analysis.

**Table S7. Cox proportional hazards models for the association between risk factor classes and incident dementia (sequential adjustment, training dataset)**

| | Age and Sex Adjusted Model | | | Fully Adjusted Model | | |
|---|---|---|---|---|---|---|
| **Variable** | **HR** | **95% CI** | **p-value** | **HR** | **95% CI** | **p-value** |
| **Age** | 1.21 | 1.20,1.23 | <0.0001 | 1.21 | 1.20,1.23 | <0.0001 |
| **Sex** | | | | | | |
| **Female** | — | — | | — | — | |
| **Male** | 1.21 | 1.13,1.28 | <0.0001 | 1.21 | 1.13,1.29 | <0.0001 |
| **Risk factor classes** | | | | | | |
| **0-1 risk factors** | — | — | | — | — | |
| **Sleep and psychosocial factors** | 1.37 | 1.23,1.52 | <0.0001 | 1.34 | 1.20,1.49 | <0.0001 |
| **Poor cardiometabolic health** | 1.38 | 1.24,1.54 | <0.0001 | 1.32 | 1.19,1.46 | <0.0001 |
| **Hearing loss and unhealthy lifestyle** | 1.39 | 1.23,1.56 | <0.0001 | 1.37 | 1.22,1.55 | <0.0001 |
| **Heavy drinking and unhealthy lifestyle** | 1.20 | 1.07,1.35 | 0.0019 | 1.20 | 1.07,1.35 | 0.0026 |
| **VitD deficiency, sedentary behaviour and psychosocial factors** | 1.20 | 1.05,1.37 | 0.0086 | 1.18 | 1.03,1.35 | 0.018 |
| **Multiple risk factors** | 2.63 | 2.33,2.96 | <0.0001 | 2.38 | 2.11 2.68 | <0.0001 |
| **Ethnicity** | | | | | | |
| **Non-White** | | | | — | — | |
| **White** | | | | 0.89 | 0.74,1.06 | 0.20 |
| **Missing** | | | | 0.59 | 0.29,1.21 | 0.15 |
| **Education** | | | | | | |
| **Primary** | | | | — | — | |
| **Secondary** | | | | 0.81 | 0.75,0.88 | <0.0001 |
| **Post-secondary non-tertiary** | | | | 0.83 | 0.75,0.91 | <0.0001 |
| **Tertiary** | | | | 0.69 | 0.64,0.76 | <0.0001 |
| **Missing** | | | | 1.20 | 0.93,1.55 | 0.17 |
| **Socioeconomic status, quintile** | | | | | | |
| **Least deprived** | | | | — | — | |
| **Second least deprived** | | | | 1.00 | 0.91,1.11 | 0.96 |
| **Middle** | | | | 1.04 | 0.95,1.15 | 0.39 |
| **Second most deprived** | | | | 1.11 | 1.01,1.22 | 0.036 |
| **Most deprived** | | | | 1.36 | 1.23,1.50 | <0.0001 |
| **Missing** | | | | 1.03 | 0.33,3.19 | >0.96 |
| **APOE-ε4 carrier status** | | | | | | |
| **non-carrier** | | | | — | — | |
| **carrier** | | | | 3.26 | 3.07,3.47 | <0.0001 |
| **Missing** | | | | 1.96 | 1.68,2.28 | <0.0001 |

Training dataset (n=121,468) was used for this analysis. HR=hazard ratio; CI=confidence interval; VitD=vitamin D; APOE=apolipoprotein E.

**Figure S3. Cox proportional hazards models for the association between risk factor classes and incident dementia by training and test dataset**

| Risk Factor Classes, by Training and test data | Cases | Total |
|---|---|---|
| **Training** | | |
| 0-1 risk factor | 593 | 22962 |
| Sleep and psychosocial factors | 811 | 23419 |
| Poor cardiometabolic health | 862 | 22468 |
| Hearing loss and unhealthy lifestyle | 509 | 13878 |
| Heavy drinking and unhealthy lifestyle | 579 | 18391 |
| VitD deficiency, sedentary and psychosocial factors | 338 | 11943 |
| Multiple risk factors | 523 | 8407 |
| **Test** | | |
| 0-1 risk factor | 171 | 5740 |
| Sleep and psychosocial factors | 177 | 5237 |
| Poor cardiometabolic health | 233 | 5396 |
| Hearing loss and unhealthy lifestyle | 117 | 3369 |
| Heavy drinking and unhealthy lifestyle | 171 | 5217 |
| VitD deficiency, sedentary and psychosocial factors | 88 | 2990 |
| Multiple risk factors | 132 | 2415 |

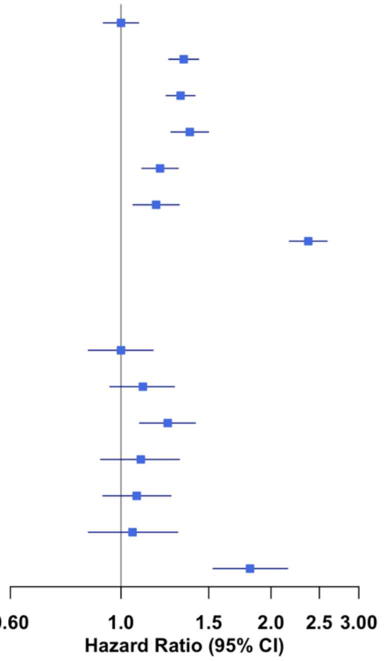


All models were adjusted for age, sex, ethnicity, education, socioeconomic status, and APOE-ε4 status. Incidence rate per 1000 person-years. Floating absolute risks are used to estimate group-specific 95% confidence intervals for all categories. Each class was characterised by risk factor patterns identified by latent class analysis. APOE=apolipoprotein E; VitD=vitamin D. The training dataset included 121,468 participants, and the test dataset included 30,364 participants.

**Table S8. Cox proportional hazards models for the association between risk factor classes and incident dementia by different follow-up periods and APOE-ε4 carrier status (training dataset)**

| Risk Factor Classes, by different subgroups | Hazard Ratio (95% CI) |
|---|---|
| **0-10 years** | |
| 0-1 risk factor | 1 (Reference) |
| Sleep and psychosocial factors | 1.26 (1.06-1.49) |
| Poor cardiometabolic health | 1.26 (1.07-1.49) |
| Hearing loss and unhealthy lifestyle | 1.42 (1.18-1.71) |
| Heavy drinking and unhealthy lifestyle | 1.18 (0.99-1.42) |
| VitD deficiency, sedentary and psychosocial factors | 1.35 (1.10-1.65) |
| Multiple risk factors | 2.51 (2.09-3.00) |
| **10- years** | |
| 0-1 risk factor | 1 (Reference) |
| Sleep and psychosocial factors | 1.39 (1.22-1.60) |
| Poor cardiometabolic health | 1.36 (1.19-1.56) |
| Hearing loss and unhealthy lifestyle | 1.34 (1.15-1.57) |
| Heavy drinking and unhealthy lifestyle | 1.21 (1.04-1.41) |
| VitD deficiency, sedentary and psychosocial factors | 1.05 (0.88-1.26) |
| Multiple risk factors | 2.27 (1.93-2.66) |
| **APOE-e4 non-carrier** | |
| 0-1 risk factor | 1 (Reference) |
| sleep and psychosocial factors | 1.48 (1.25-1.75) |
| poor cardiometabolic health | 1.60 (1.36-1.88) |
| hearing loss and unhealthy lifestyle | 1.58 (1.32-1.90) |
| heavy drinking and unhealthy lifestyle | 1.42 (1.19-1.70) |
| VitD deficiency, sedentary and psychosocial factors | 1.25 (1.02-1.54) |
| multiple risk factors | 3.55 (2.93-4.22) |
| **APOE-e4 carrier** | |
| 0-1 risk factor | 1 (Reference) |
| Sleep and psychosocial factors | 1.25 (1.08-1.44) |
| Poor cardiometabolic health | 1.14 (0.98-1.31) |
| Hearing loss and unhealthy lifestyle | 1.25 (1.06-1.47) |
| Heavy drinking and unhealthy lifestyle | 1.04 (0.89-1.23) |
| VitD deficiency, sedentary and psychosocial factors | 1.15 (0.96-1.38) |
| Multiple risk factors | 1.59 (1.33-1.90) |

Models by different follow-up periods were adjusted for age, sex, ethnicity, education, socioeconomic status, and APOE-ε4 carrier status. Models by APOE-ε4 carrier status were adjusted for age, sex, ethnicity, education, and socioeconomic status. Each class was characterised by risk factor patterns identified by latent class analysis. Training dataset (n=121,468) was used for this analysis. APOE=apolipoprotein E; VitD=vitamin D.

**Table S9. Cox proportional hazards models for the association between risk factor classes and incident dementia by sex (training dataset)**

| Risk Factor Classes, by different subgroups | Hazard Ratio (95% CI) |
|---|---|
| **Male** | |
| 0-1 risk factor | 1 (Reference) |
| Sleep and psychosocial factors | 1.30 (1.10-1.52) |
| Poor cardiometabolic health | 1.31 (1.12-1.52) |
| Hearing loss and unhealthy lifestyle | 1.48 (1.25-1.74) |
| Heavy drinking and unhealthy lifestyle | 1.18 (1.01-1.38) |
| VitD deficiency, sedentary and psychosocial factors | 1.24 (1.02-1.51) |
| Multiple risk factors | 2.41 (2.04-2.84) |
| **Female** | |
| 0-1 risk factor | 1 (Reference) |
| Sleep and psychosocial factors | 1.37 (1.19-1.58) |
| Poor cardiometabolic health | 1.33 (1.15-1.54) |
| Hearing loss and unhealthy lifestyle | 1.24 (1.04-1.48) |
| Heavy drinking and unhealthy lifestyle | 1.30 (1.05-1.60) |
| VitD deficiency, sedentary and psychosocial factors | 1.12 (0.94-1.35) |
| Multiple risk factors | 2.34 (1.96-2.78) |

Models were adjusted for age, ethnicity, education, socioeconomic status, and APOE-ε4 carrier status. P-values for interaction between sex and risk factor classes: Sleep and psychosocial factors p=0.61; Poor cardiometabolic health p=0.84; Hearing loss and unhealthy lifestyle p=0.17; Heavy drinking and unhealthy lifestyle p=0.49; Vitamin D deficiency, sedentary and psychosocial factors p=0.48; Multiple risk factors p=0.81. Training dataset (n=121,468) was used for this analysis.

**Table S10. Cox proportional hazards models for the association between risk factor classes and incident dementia using attained age as the underlying time scale (training dataset)**

| **Risk Factor Classes** | **Hazard Ratio (95% CI)** |
|---|---|
| 0-1 risk factor | 1 (Reference) |
| Sleep and psychosocial factors | 1.33 (1.20-1.48) |
| Poor cardiometabolic health | 1.30 (1.17-1.44) |
| Hearing loss and unhealthy lifestyle | 1.37 (1.21-1.54) |
| Heavy drinking and unhealthy lifestyle | 1.20 (1.07-1.35) |
| VitD deficiency, sedentary and psychosocial factors | 1.18 (1.04-1.35) |
| Multiple risk factors | 2.36 (2.09-2.66) |

Models were adjusted for sex, ethnicity, education, socioeconomic status, and APOE-ε4 carrier status. Attained age was used as the underlying time scale. Training dataset (n=121,468) was used for this analysis.

**Table S11. Cox proportional hazards models for the association between risk factor classes and incident dementia (excluding prevalent cardiovascular disease, training dataset)**

| Risk Factor Classes | Hazard Ratio (95% CI) |
|---|---|
| 0-1 risk factor | 1 (Reference) |
| Sleep and psychosocial factors | 1.28 (1.15-1.44) |
| Poor cardiometabolic health | 1.22 (1.09-1.37) |
| Hearing loss and unhealthy lifestyle | 1.32 (1.16-1.50) |
| Heavy drinking and unhealthy lifestyle | 1.20 (1.06-1.36) |
| VitD deficiency, sedentary and psychosocial factors | 1.17 (1.02-1.35) |
| Multiple risk factors | 2.09 (1.82-2.40) |

Models were adjusted for age, sex, ethnicity, education, socioeconomic status, and APOE-ε4 carrier status. Participants with prevalent cardiovascular disease (CVD) at baseline (n=12,331, 10.2% of the training dataset) were excluded, leaving 109,137 participants for analysis. CVD was defined as self-reported history of coronary heart disease, heart failure, cerebrovascular disease, or aortic and peripheral arterial disease. Training dataset was used for this analysis.

**Table S12. Cox proportional hazards models for the association between risk factor classes and incident dementia (0 risk factor reference, training dataset)**

| Risk Factor Classes | Hazard Ratio (95% CI) |
|---|---|
| 0 risk factor | 1 (Reference) |
| 1 risk factor | 1.17 (0.96-1.43) |
| Sleep and psychosocial factors | 1.51 (1.25-1.83) |
| Poor cardiometabolic health | 1.49 (1.23-1.80) |
| Hearing loss and unhealthy lifestyle | 1.55 (1.27-1.89) |
| Heavy drinking and unhealthy lifestyle | 1.35 (1.11-1.65) |
| VitD deficiency, sedentary and psychosocial factors | 1.33 (1.08-1.64) |
| Multiple risk factors | 2.69 (2.20-3.28) |

Models were adjusted for age, sex, ethnicity, education, socioeconomic status, and APOE-ε4 carrier status. Among the participants with 0-1 risk factors (n=22,962), 5,449 participants had 0 risk factor and 17,513 participants had 1 risk factor. Training dataset (n=121,468) was used for this analysis.
